# Public health interventions successfully mitigated multiple incursions of SARS-CoV-2 Delta variant in the Australian Capital Territory

**DOI:** 10.1101/2022.08.23.22278828

**Authors:** Robyn Hall, Ashley Jones, Emma Crean, Victoria Marriott, Nevada Pingault, Alexandra Marmor, Timothy Sloan-Gardner, Karina Kennedy, Kerryn Coleman, Vanessa Johnston, Benjamin Schwessinger

## Abstract

The Australian Capital Territory rapidly responded to an incursion of the SARS-CoV-2 Delta (B.1.617.2) variant on 12 August 2021 with several public health interventions, including a territory-wide lockdown and genomic sequencing. Prior to this date, SARS-CoV-2 had been eliminated locally since July 7, 2020. Sequencing of >75% of cases identified at least 13 independent incursions with onwards spread in the community during the study period, between 12 August and 11 November 2021. Two incursions resulted in the majority of community transmission during this period, with persistent transmission in vulnerable sections of the community. Ultimately, both major incursions were successfully mitigated through public health interventions, including COVID-19 vaccines. In this study we explore the demographic factors that contributed to the spread of these incursions. The high rates of SARS-CoV-2 sequencing in the Australian Capital Territory and the relatively small population size facilitated detailed investigations of the patterns of virus transmission. Genomic sequencing was critical to disentangling complex transmission chains to target interventions appropriately.

- Despite a strict lockdown and interstate travel restrictions, the Australian Capital Territory experienced at least 13 incursions of SARS-CoV-2 Delta (B.1.617.2) with onwards spread in the community between 12 August and 11 November 2021.
- This level of detail was only accessible because of the high rate of SARS-CoV-2 sequencing, with sequencing attempted on 1438/1793 (80%) of cases.
- Transmission chains varied in size and duration, with two dominant incursions (ACT.19 and ACT.20) comprising 35% and 53% of all sequenced cases during the study period, respectively.
- The ACT.20 outbreak persisted longer, due to specific challenges with implementing public health interventions in the affected populations.
- Both major incursions were successfully curbed through stringent public health measures, including the widespread acceptance of COVID-19 vaccines (>95% of the eligible population by the end of the study period).

## Introduction

After the emergence of SARS-CoV-2 in late 2019 in Asia, the Australian Government declared a human biosecurity emergency on 18 March 2020 and closed its international borders to non-permanent residents and non-citizens on 20 March 2020 [1]. All inbound passengers were required to undertake two weeks of supervised hotel quarantine with mandatory testing [1]. This largely restricted the impacts of the COVID-19 pandemic during 2020 and 2021, compared to much of the rest of the world. Australia experienced a first wave of local SARS-CoV-2 transmission, with the so-called ‘ancestral’ SARS-CoV-2 variant, from March to April of 2020, predominantly driven by returning overseas travellers and cruise ship passengers [2]. A second wave, again due to ‘ancestral’ SARS-CoV-2, was experienced in June to October of 2020, primarily in the south-eastern state of Victoria (VIC) with spread to other jurisdictions before lockdowns were enforced [3]. Notably, household transmission associated with three returning travellers from Victoria was detected in the ACT linked to this outbreak [4]. A third wave, due to the B.1.617.2 Delta variant of concern, began in New South Wales (NSW) on 15 June 2021 after an unvaccinated limousine driver was infected while transporting international air crew [5]. This wave subsequently spread to all other Australian jurisdictions. While minor incursions, rapidly controlled through public health interventions, were experienced in Western Australia (WA), South Australia (SA), the Northern Territory (NT), Queensland (QLD), and Tasmania (TAS), prolonged community transmission occurred in NSW, VIC, and the Australian Capital Territory (ACT) [6]. With increasing population vaccination coverage, most domestic restrictions were lifted in November 2021, including international travel for vaccinated citizens and permanent residents, and Delta was replaced by the B.1.1.529 Omicron variant in December 2021 [7].

The ACT is a small (2358 km^2^) enclave within NSW in south-eastern Australia with a population of approximately 453,558 people. During the first and second waves, the ACT experienced relatively little community transmission of SARS-CoV-2 (29 cases to 3 January 2021) [8]; the last local transmission prior to the Delta outbreak occurred on July 7 2020 [9]. There were no local restrictions (i.e., density limits, mask wearing) during the first half of 2021 and an elimination strategy with a strong focus on ‘trace, test, isolate, and quarantine’ was in place pending the rollout of vaccines to the eligible population. Approved vaccines (Vaxzevria (AstraZeneca), Comirnaty (Pfizer), and later Spikevax (Moderna)) became available from 22 February 2021 via a phased approach, initially targeting frontline workers and high-risk individuals [10]. However, by 15 June, when the third wave started in NSW, vaccines were still not available for much of the general population, with Comirnaty restricted primarily to individuals aged 40 years and older. Only on 29 June 2021 was Vaxzevria approved for use in younger individuals; this vaccine was previously restricted to older age groups due to concerns around thrombosis with thrombocytopenia syndrome in younger persons [11]. Comirnaty became available in the ACT to those 30 years and older on 3 August, those 16 years and older from 1 September, and those 12 years and older from 13 September. On 23 June the ACT government implemented interstate travel restrictions for travellers from high-risk areas. Masks became compulsory in indoor settings from 28 June, but these restrictions were lifted on 10 July. A QR code check-in application was made mandatory for most businesses from 15 July to assist in contact tracing efforts in the event of an incursion.

On 12 August 2021, 398 days after the last local transmission of SARS-CoV-2 in the ACT, a case was detected in an individual with no known travel history to NSW, triggering a local lockdown and strict enforcement of mask mandates and use of the QR code check-in application (Figure 1). All cases underwent contact tracing, and cases and close contacts (defined as (*i*) a member of the same household, or (*ii*) a person notified by an authorised person that they were a close contact) were required to isolate or quarantine, respectively, for 14 days. Full genome sequencing of SARS-CoV-2 was attempted for most cases, to assist with contact tracing efforts by identifying discordant transmission pathways and identifying new incursion events. Lockdown was lifted on 15 October, due to high vaccination coverage and decreasing case counts (Figure 1) [12]. On-campus learning for schools returned in a staged approach; year 12 students returned on 5 October, while other year groups and early childhood services returned between 18 October and 1 November. Remaining restrictions such as density limits and mask requirements (in most settings) were lifted on 12 November; at this time, 95% of ACT residents aged 12 and over (81% of the total population) had received two COVID-19 vaccinations. This was the first application of real-time genomic epidemiology at the local level in the ACT. Here we describe the utility of genomic epidemiology during the SARS-CoV-2 Delta outbreak in this jurisdiction in conjunction with traditional contact tracing efforts and prior to the lifting of public health interventions.

**Figure 1:**
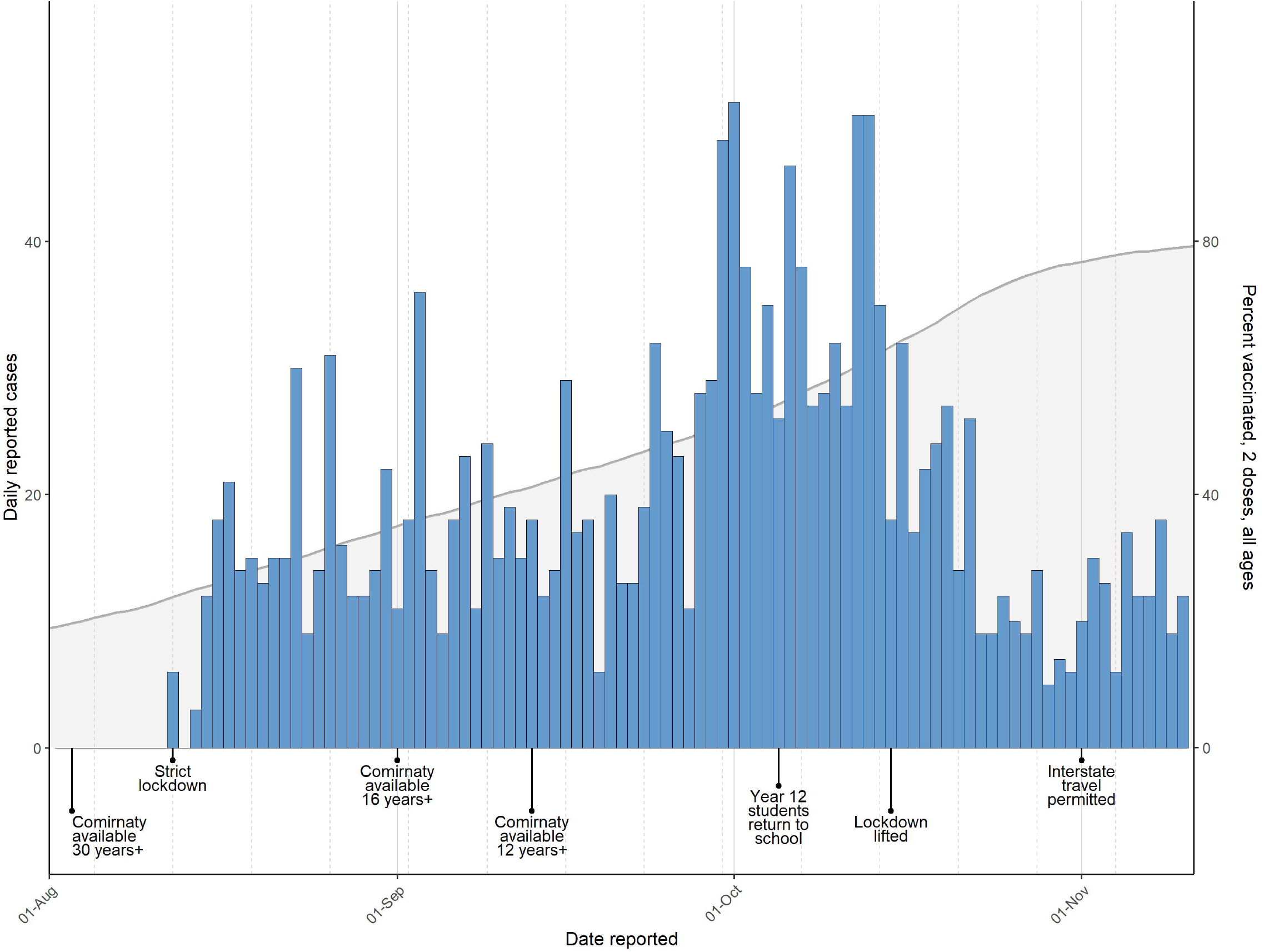
Epidemic curve and timeline of public health interventions for SARS-CoV-2 in the Australian Capital Territory, August to November 2021. The number of reported cases (left y-axis) per day is shown as a bar chart. The cumulative percentage of the total population receiving two vaccine doses (right y-axis) is shown as a grey area curve. A timeline of the major public health interventions is shown below the charts.

## Methods

### Sampling

For this study, cases were restricted to the 3-month period between 12 August 2021 and 11 November 2021, when public health interventions were in place. All laboratory-confirmed SARS-CoV-2 cases in the ACT were notified to ACT Health, and core and enhanced epidemiological, demographic, and clinical data were collected through telephone interviews using a standard questionnaire. Ethnicity information for positive cases was self-reported based on the question “How would you describe your ethnic or cultural background?” using the Australian Bureau of Statistics Australian Standard Classification of Cultural and Ethnic Groups. Vaccination data were obtained from ACT Health and population denominators were obtained from the Australian Bureau of Statistics. Positive RNA extracts from the two major local testing laboratories, ACT Pathology and Capital Pathology, were forwarded for pathogen sequencing at the Australian National University. We were not able to obtain specimens for some cases that were tested in other jurisdictions. Conversely, we received 30 specimens that were not classified as ACT cases but were included in our analyses. From 29 September we did not attempt sequencing of samples with RT-qPCR cycle thresholds ≥33 or from cases that were household contacts of a sequenced case.

### SARS-CoV-2 sequencing

SARS-CoV-2 sequencing was performed using the ARTIC Network amplicon sequencing protocol, using either Superscript IV (Life Technologies) or Lunascript (New England Biolabs) reverse transcriptase for cDNA synthesis [13]. For PCR amplification, we initially used the v3 primer set until 10 October 2021, when we switched to v4 [14]. Pooled amplicon libraries were prepared using the Ligation Sequencing Kit (Oxford Nanopore Technologies (ONT) EXP-AMII001) and native barcodes (ONT EXP-NBD196). Sequencing was performed on a MinION Mk1B using a R9.4.1 FLO-MIN106D flow cell, as per manufacturer’s instructions (ONT). Sequencing progress was monitored with RAMPART [15] until sufficient coverage was achieved. Raw fast5 sequencing reads were basecalled to fastq and demultiplexed using Guppy (versions 4 and 5, ONT). Consensus sequences were generated using the ARTIC network SARS-CoV-2 bioinformatics pipeline version 1.2.1 against the MN908947.3 reference sequence [16]. EPI_ISL_3643665 was processed and sequenced using Illumina technology at the Microbial Genomics Reference Laboratory, NSW Health Pathology (NSWHP) Institute of Clinical Pathology and Medical Research. EPI_ISL_5587718 was processed and sequenced using the Oxford Nanopore MinION at NSWHP Royal Prince Alfred Hospital.

Sequencing turnaround time was estimated by calculating the time from sample collection to preliminary reporting of the sequencing results to ACT Health. Since no time stamp was available for sample collection, we used 12:00.

### Phylogenetic analysis

Sequences with ≤20% ambiguous bases were aligned against the Wuhan-1 (GenBank Accession NC_045512 [17]) reference sequence using the FFT-NS-2 algorithm in MAFFTv7.450 [18] as implemented in Geneious Prime 2021.2.2 (https://www.geneious.com/). The alignment was curated manually to check for gaps and misalignments. Sequences were assigned to ACT genomic lineages and sublineages based on neighbor-joining phylogenies (≤10% ambiguous bases only) constructed using the Tamura-Nei model as implemented in the Geneious Tree Builder in Geneious Prime 2021.2.2 and/or on manual exploration of the alignments at lineage-defining sites. Lineage assignment was revised daily during the COVID-19 response, based on the incorporation of new sequences and in conjunction with epidemiological information. Comparisons to publicly available Australian and international sequences were performed with the UShER webserver [19].

To estimate a time-scaled phylogeny, a maximum likelihood (ML) phylogeny was estimated using iqtreev2.1.2 [20] for sequences with ≤10% ambiguous bases (n = 1275). Branch support was estimated using 1000 ultrafast bootstrap approximations [21] and 1000 replicates of the SH-like approximate likelihood ratio test [22]. This ML tree, along with sample collection dates, were used as input for treetimev0.8.5 [23]. The tree was rooted on NC_045512 (https://www.ncbi.nlm.nih.gov/genbank/).

Figures were generated in Rv4.1.0 using the following packages: tidyversev1.3.1 [24], ggtreev3.3.0.900 [25], scalesv1.1.1 [26], ggaltv0.4.0 [27], and cowplotv1.1.1 [28].

## Results

From the 12 August 2021 to 11 November 2021, 1793 laboratory-confirmed SARS-CoV-2 infections were reported in ACT residents. Of these, SARS-CoV-2 sequencing was attempted for 1438 cases (80%). Near-complete genomes (≤1% ambiguous bases) were recovered from 287 cases (16.0% of all ACT cases), we recovered 960 partial genomes (1– ≤10% ambiguous bases; 53.5% of all ACT cases), and 100 poor quality genomes (10–≤20% ambiguous bases; 5.6% of all ACT cases). The remaining 91 cases for which sequencing was attempted yielded incomplete genomes (>20% ambiguous bases). The estimated turnaround time for sequences and analyses to become available for public health action was within 3.2 days for 50% of sequences and within 7.0 days for 95% of sequences. Additionally, we sequenced 30 samples from non-ACT cases that were received through ACT laboratories.

Based on the phylogeny and corroborating epidemiological data, we identified at least 13 incursions into the ACT resulting in forward transmission in the community over the study period, despite interstate travel restrictions. Each incursion was classified as a separate ACT genomic lineage (Figure 2). Importantly, this sets a lower bound to the number of possible incursions, since the genetic diversity of SARS-CoV-2 circulating in Australia was limited at this time, therefore multiple incursions of near-identical SARS-CoV-2 genomes would likely have been missed. Furthermore, we identified 13 sequences as genomic singletons that did not cluster (≤2 nucleotide differences) with another ACT sequence, and three lineages that were contained to a single household. These introductions did not result in forward transmission within the community and were likely contained through the strict 14-day quarantine restrictions for returning residents.

**Figure 2:**
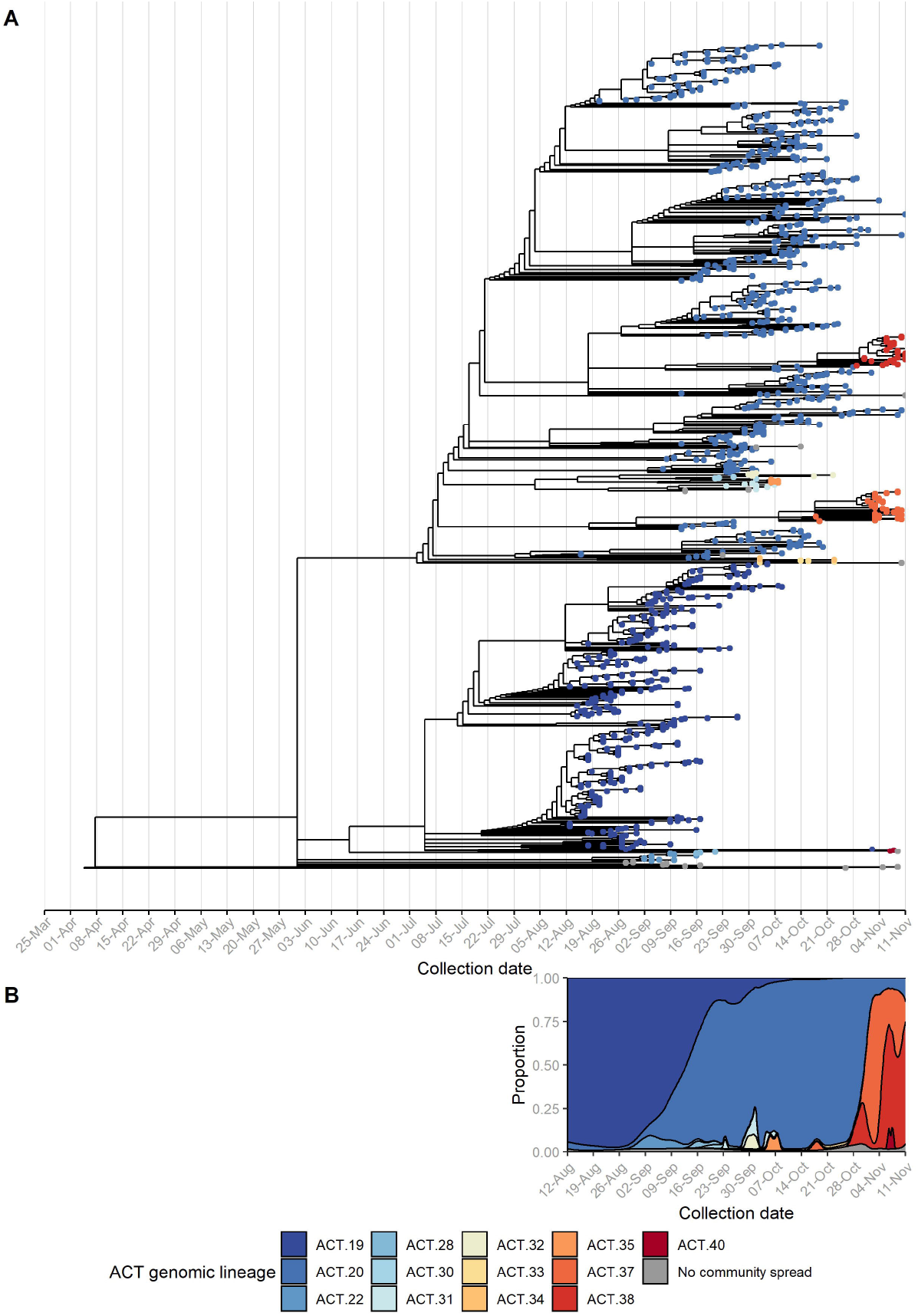
Incursions and onward spread of SARS-CoV-2 B.1.617.2 (Delta) in the Australian Capital Territory (ACT), 12 August to 11 November 2021. Sequencing of SARS-CoV-2 was attempted on 80% of ACT cases reported during the study period and an additional 30 non-ACT cases. A time-structured phylogeny was estimated based on consensus sequences with ≤10% ambiguous bases (A). Tips are coloured by ACT genomic lineage. Each lineage reflects a separate incursion event with subsequent local spread, as defined by phylogenetic analysis and corroborating epidemiological information. Sequences where onwards transmission within the community was not identified within the ACT are coloured grey. The density plot shows the relative proportion of ACT genomic lineages over time, based on all sequences with ≤20% ambiguous bases (B). Both A) and B) are scaled to the same x-axis.

The individual incursions led to ongoing transmission chains that varied considerably in size, duration, and demographics (Table 1). The Delta (B.1.617.2) “wave” in the ACT was dominated by two large incursions, ACT.19 and ACT.20, both initially identified early in the outbreak (12 August and 19 August, respectively). By mid-September, the ACT.19 incursion had been mostly controlled through timely and effective public health interventions, such as a territory-wide lockdown and mask mandates, at which point ACT.20 was detected more widely. ACT.20 case numbers declined throughout October, which negatively correlated with the number of vaccine doses delivered (Figure 1) [12]. However, sporadic cases continued to be identified through to the end of the study period.

**Table 1:**
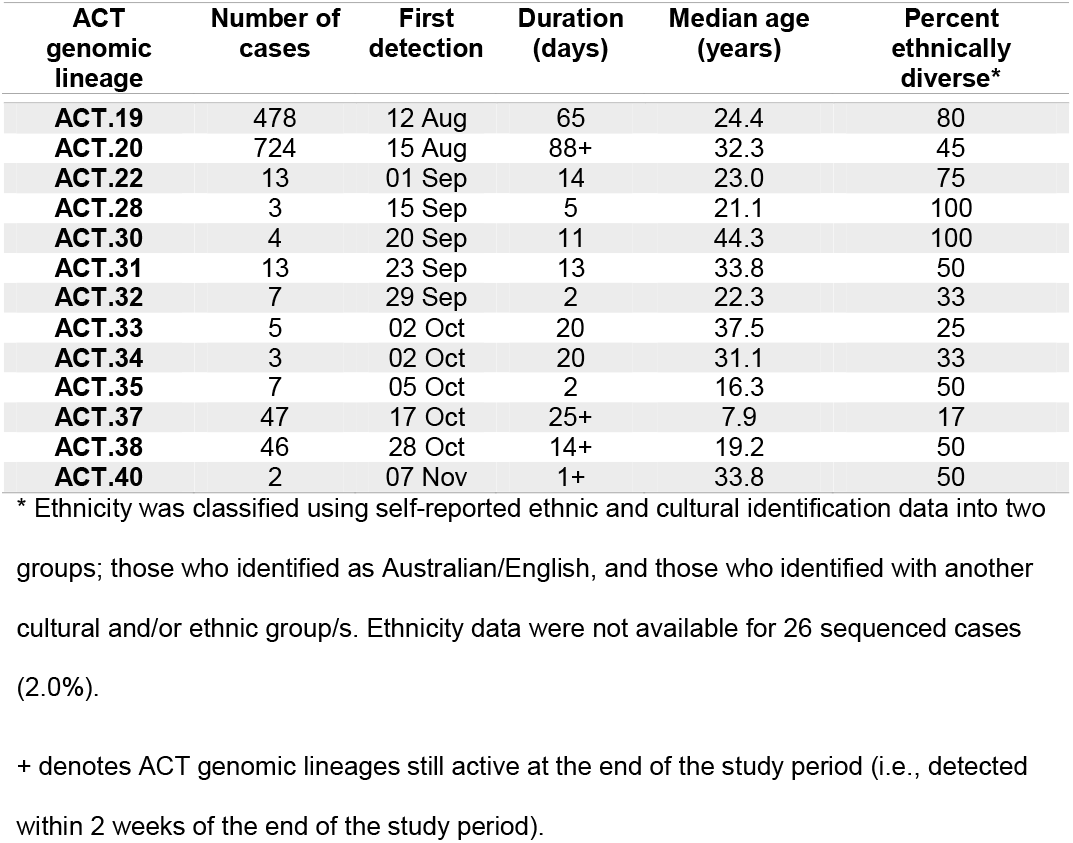
Size, duration, and epidemiological characteristics of SARS-CoV-2 B.1.617.2 (Delta) incursions.

| ACT genomic lineage | Number of cases | First detection | Duration (days) | Median age (years) | Percent ethnically diverse* |
| --- | --- | --- | --- | --- | --- |
| **ACT.19** | 478 | 12 Aug | 65 | 24.4 | 80 |
| **ACT.20** | 724 | 15 Aug | 88+ | 32.3 | 45 |
| **ACT.22** | 13 | 01 Sep | 14 | 23.0 | 75 |
| **ACT.28** | 3 | 15 Sep | 5 | 21.1 | 100 |
| **ACT.30** | 4 | 20 Sep | 11 | 44.3 | 100 |
| **ACT.31** | 13 | 23 Sep | 13 | 33.8 | 50 |
| **ACT.32** | 7 | 29 Sep | 2 | 22.3 | 33 |
| **ACT.33** | 5 | 02 Oct | 20 | 37.5 | 25 |
| **ACT.34** | 3 | 02 Oct | 20 | 31.1 | 33 |
| **ACT.35** | 7 | 05 Oct | 2 | 16.3 | 50 |
| **ACT.37** | 47 | 17 Oct | 25+ | 7.9 | 17 |
| **ACT.38** | 46 | 28 Oct | 14+ | 19.2 | 50 |
| **ACT.40** | 2 | 07 Nov | 1+ | 33.8 | 50 |

\* Ethnicity was classified using self-reported ethnic and cultural identification data into two groups; those who identified as Australian/English, and those who identified with another cultural and/or ethnic group/s. Ethnicity data were not available for 26 sequenced cases (2.0%).
+ denotes ACT genomic lineages still active at the end of the study period (i.e., detected within 2 weeks of the end of the study period).

Both incursions particularly impacted vulnerable groups within the community where crowded living arrangements and lower health literacy were contributing factors; however, the specific demographic characteristics of these vulnerable populations were distinct in the two outbreaks. To assist with active case finding and control efforts, a mobile (in-reach) testing and vaccination strategy was rolled out by partnering with health services and non-government organizations.

Cases related to the ACT.19 incursion frequently were from large ethnically diverse households with non-nuclear families, often spanning multiple residences (Table 1). For comparison, while the Australian Bureau of Statistics does not report ethnicity data directly, in the 2021 national census 24.6% of ACT persons in the ACT reported speaking a language other than English at home and 29% reported being born overseas. The number of people per household was relatively high and community engagement was complex (Supplementary figure 1). Additionally, many were essential workers, often with carer responsibilities, resulting in forward transmission into other vulnerable populations. To assist with control efforts, ACT Health engaged extensively with community and cultural leaders, cross-government and non-government agencies, and focussed efforts on the provision of culturally appropriate and in-reach supports.

In contrast, the ACT.20 incursion predominantly affected a less ethnically diverse and more socially disadvantaged cohort (Table 1). While there were often fewer people per household in this group (Supplementary figure 1), there were challenges around contact tracing and access to testing.

The phase after the lifting of the territory-wide lockdown (on October 15) was characterized by two medium-sized incursions, ACT.37 and ACT.38. The median age of cases in these incursions was eight and 19 years, respectively (Table 1); this was significantly lower than the median age of non-ACT.37 or ACT.38 cases (i.e., all cases belonging to other ACT genomic lineages or genomic singletons), which was 28 years (Mood’s test, adjusted *p* = 5e^-3^ and 2e^-5^, respectively). Of relevance, schools reopened between 5 October and 1 November and vaccines were not available for those aged 16 to 18 years until 1 September, and for those aged 12 to 16 years until 13 September. Contact tracing revealed that one of these incursions was linked to a school setting [29], while the other was attributed to a party at a private residence [30]. Genomic sequencing showed that the school-associated incursion was limited to students and their immediate contacts (e.g., parents, siblings, and other household members); there was no extended community transmission related to this incursion. In contrast, the spread from the private party was more extensive.

By 11 November (the end of the study period) all incursions were considered to be sufficiently controlled and population vaccination coverage was high, leading to the lifting of most restrictions. Notably, during the study period there were 11 COVID-19-related deaths. The extensive contact tracing and case follow-up employed in the ACT likely facilitated the early identification of those cases eligible for enhanced treatment, such as monoclonal antibody therapies. The high vaccination rates achieved in the ACT and good compliance with public health social measures during the outbreak period likely contributed to the low observed mortality [12].

In addition to the tracking of broad-scale ACT genomic lineages, we used single mutations to define genomic sublineages. These were found to map closely to epidemiologically defined case clusters and this sublineage information was used to link cases with an unknown source of acquisition to clusters and to resolve complex transmission chains, such as where cases were linked to multiple exposure locations. For example, genomic sequencing revealed links between two different high schools via common exposure through extra-curricular activities (Figure 3) [31].

**Figure 3:**
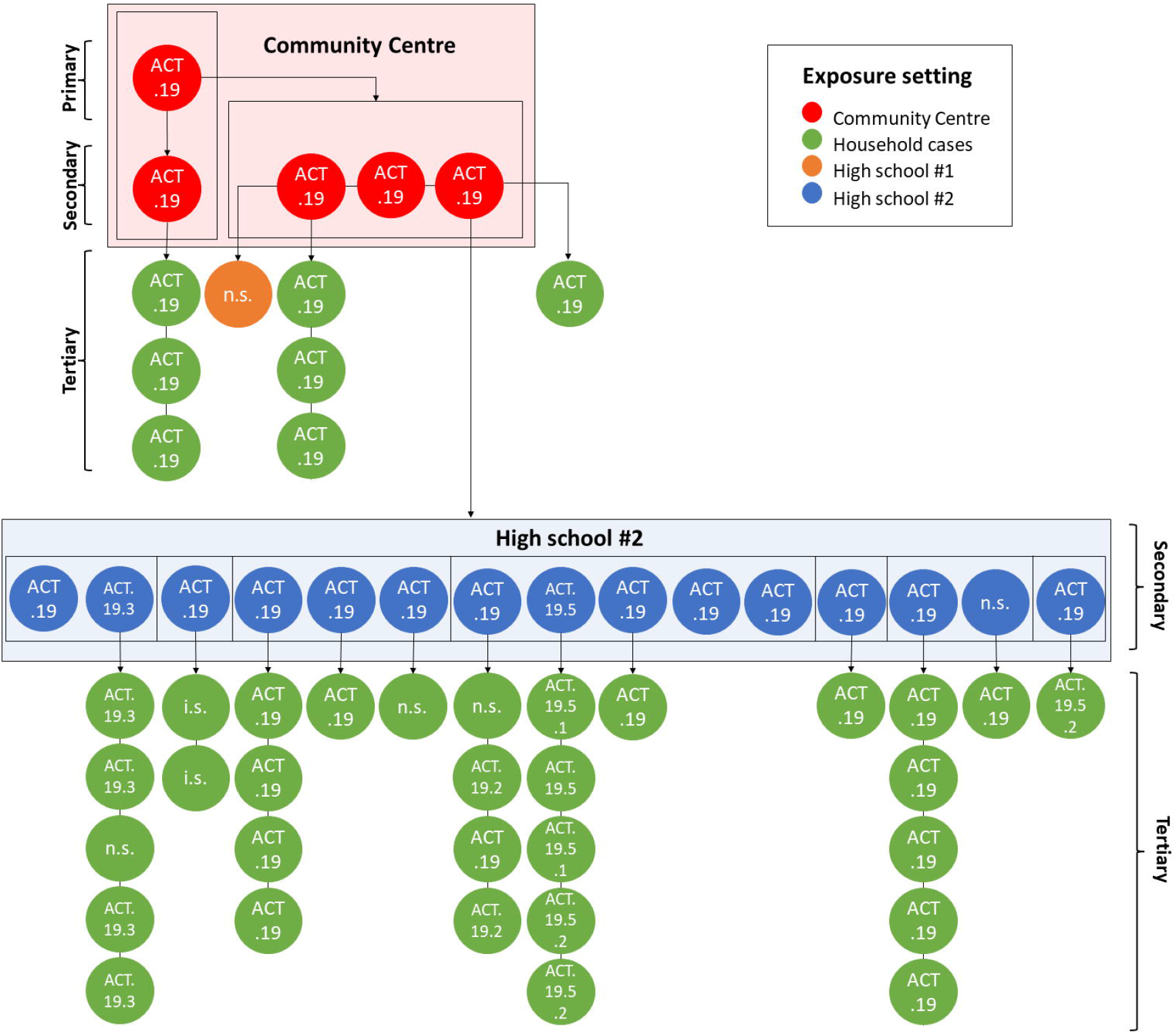
Resolution of a complex transmission chain using genomic epidemiology. Dots represent individual cases and are coloured by exposure setting. Primary, secondary, and tertiary cases are marked by braces. The ACT genomic sublineage of each case is specified. Directionality of transmission, inferred from epidemiological contact tracing, laboratory information, and/or genomic sequencing, is indicated by arrows. Boxes delineate separate cohorts. n.s. not sequenced, i.s. case diagnosed interstate.

Incorporating this genomic contact tracing, we were able to identify transmission at certain exposure sites and implement enhanced infection control measures in these settings. For example, genomic information showed that a case cluster at a social housing complex was the result of several incursions over a 3-week period, rather than a single superspreading event which was assumed based only on case interviews and contact tracing information (Figure 4). While isolated cases were identified in other high-risk settings, such as hospitals and correctional facilities, genomic sequencing showed that typically these cases were community-acquired and that in most of these settings (apart from a single outbreak in a residential aged care facility) there was no significant spread. Together, these demonstrate some examples of how genomic sequencing was used in the ACT to inform outbreak mitigation strategies.

**Figure 4:**
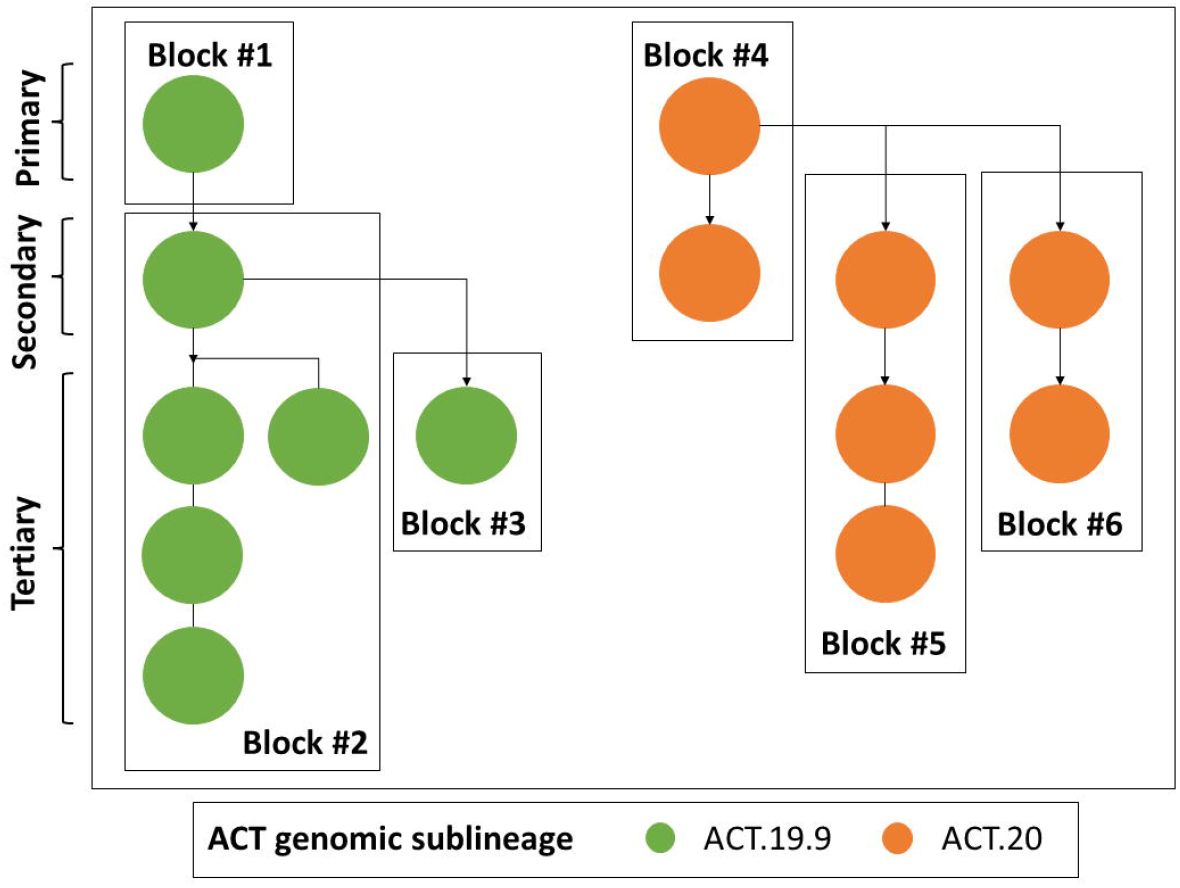
Genomic sequencing revealed two independent incursions into a social housing complex over a 3-week period. Dots represent individual cases and are coloured by ACT genomic sublineage. Primary, secondary, and tertiary cases are marked by braces. Directionality of transmission, inferred from epidemiological contact tracing, laboratory information, and/or genomic sequencing, is indicated by arrows. Boxes delineate separate cohorts.

## Discussion

The combination of stringent and timely public health interventions, exhaustive contact tracing (due to the relatively small population size), and high levels of viral sequencing effectively limited the two major Delta incursions, ACT.19 and ACT.20, within a 3-month period. A key feature of the Delta (B.1.617.2) SARS-CoV-2 outbreak in the ACT was the very high proportion of cases for which sequencing was attempted (>80% of all reported cases), leading to an unprecedented understanding of local transmission networks and number of distinct incursions. The rapid turnaround time for sequencing results (< 7.0 days in 95% of cases and < 3.2 days in 50% of cases) greatly facilitated epidemiological contact tracing and the identification of high-risk exposure locations. Notably, the two dominant incursions during the resolution of ACT.20 (i.e., ACT.37 and ACT.38) disproportionately affected incompletely vaccinated younger persons, with transmission occurring at (*i*) a large private party, and (*ii*) within a school setting, demonstrating the marked impacts of even superspreading events. The cohorting strategy applied in the school setting as students and teachers returned to on-campus learning, in addition to other COVID-safe measures such as masking, was very effective in limiting the extent of community transmission associated with this incursion.

The two major incursions, ACT.19 and ACT.20, varied considerably in the affected demographic groups, and subsequently in duration and total number of cases. Several studies have already demonstrated a disproportionate risk of SARS-CoV-2 infection in socioeconomically disadvantaged groups and in ethnic minorities [32-36], and it is well-established that socioeconomic factors are generally important determinants of health and disease. At a local level, the use of genomic epidemiology to differentiate between the two overlapping incursions led to a greater understanding of transmission in at-risk populations and the need for enhanced mitigation measures in these settings, such as culturally appropriate engagement and the deployment of in-reach interventions in association with non-government organizations.

Interestingly, the first cases of ACT.20 were detected on 15 August 2021, three days after the detection of ACT.19. Yet cases only began to increase exponentially in early September and there was very limited genetic diversity in the month following detection, with the first mutation detected in samples collected on 11 September 2021. It is possible that there was undetected transmission of ACT.20 after the first detection, although testing rates were high, there was exhaustive contact tracing of cases, and strict lockdowns were in place. Alternatively, there may have been a separate introduction of a genetically identical virus. While this may seem unlikely, the diversity of SARS-CoV-2 in Australia during mid-2021 was limited, as all local cases arose from a single point source outbreak in Sydney, NSW, on 15 June 2020 [5]. Indeed, of the publicly available Australian sequences, over 400 of these are identical to the ACT.20 sequence, and it is very likely that many more cases were either not detected, not successfully sequenced, or not uploaded. This highlights the challenges around identifying individual incursions early in an outbreak when genetic diversity is limited [37-39].

While we observed complete replacement of ACT.19 by ACT.20, and near-complete replacement of ACT.20 by ACT.37 and ACT.38, these replacements were not a consequence of enhanced epidemiological fitness of any of these viruses. Indeed, the ACT.20 founder sequence had only five non-synonymous changes relative to the ACT.19 founder sequence across all coding sequences, only one of which was in the spike protein, while ACT.37 and ACT.38 each had two non-synonymous changes relative to ACT.20, none of which were in the spike protein. The spread of SARS-CoV-2 Delta in the ACT is a clear example of repeated founder effects, because of the extensive mitigation measures and the stochastic nature of transmission of SARS-CoV-2 (overdispersion) [40]. This could only be revealed through high levels of genomic sequencing in this relatively small population.

We describe here the progression of the SARS-CoV-2 Delta incursion in the ACT, highlighting the utility of intensive genomic sequencing in the public health response. We show that the Delta outbreak was driven by several independent incursions, with successive waves impacting different vulnerable groups. However, by deploying enhanced interventions (such as culturally appropriate engagement and partnered in-reach interventions) into these at-risk communities, timely public health measures were successful in mitigating these incursions and most importantly, in preventing severe clinical outcomes.

## Data Availability

All sequences are available in GISAID. Accession IDs are provided in Supplementary table 1.

https://gisaid.org/

## Acknowledgements

We gratefully acknowledge all current and previous members of the SARS-CoV-2 Schwessinger lab sequencing team: Austin Bird, Carolina Correa Ospina, Bayantes Dagvadorj, Scott Ferguson, Abigail Graetz, Evie Hodgson, Elise Kellett, Rachel Leonard, Carl McCombe, Lydia Murphy, Catalina Barragán Quintero, Rene Riedelbauch, Makenna Short, Gabrielle Smith, Rita Tam, Salome Wilson and Daniel Yu. We kindly thank the Microbial Genomics Reference Laboratory, NSW Health Pathology - Institute of Clinical Pathology and Medical Research for provision of sequencing data for EPI_ISL_3643665. We kindly thank New South Wales Health Pathology -Royal Prince Alfred Hospital for providing sequence EPI_ISL_5587718. We thank the laboratory teams at ACT Pathology and Capital Pathology, notably Craig Kennedy, Sandra Molloy, and Paul Whiting, for providing the RNA extracts for sequencing. This work and the associated public health impact was a truly cooperative effort with the entire ACT Health COVID-19 team, particularly the case investigation and epidemiology teams.

## Data Availability Statement

All sequences are available in GISAID. Accession IDs are provided in Supplementary table 1.

## Ethics statement

This work was conducted as part of the public health response to COVID-19 under the *Public Health Act 1997*. As such, no ethics approvals were required.

## Figure legends

**Supplementary Figure 1: Number of cases per household as a proxy of household size.** For the ACT.19 and ACT.20 genomic lineages, the percent of households with *n* cases per household is shown. Household contact data were not available for 88 and 360 sequenced cases for ACT.19 and ACT.20, respectively (20% and 54%).

