## Supplementary figures and images for "Public health interventions successfully mitigated multiple incursions of SARS-CoV-2 Delta variant in the Australian Capital Territory"

### Supplemental Figure 1

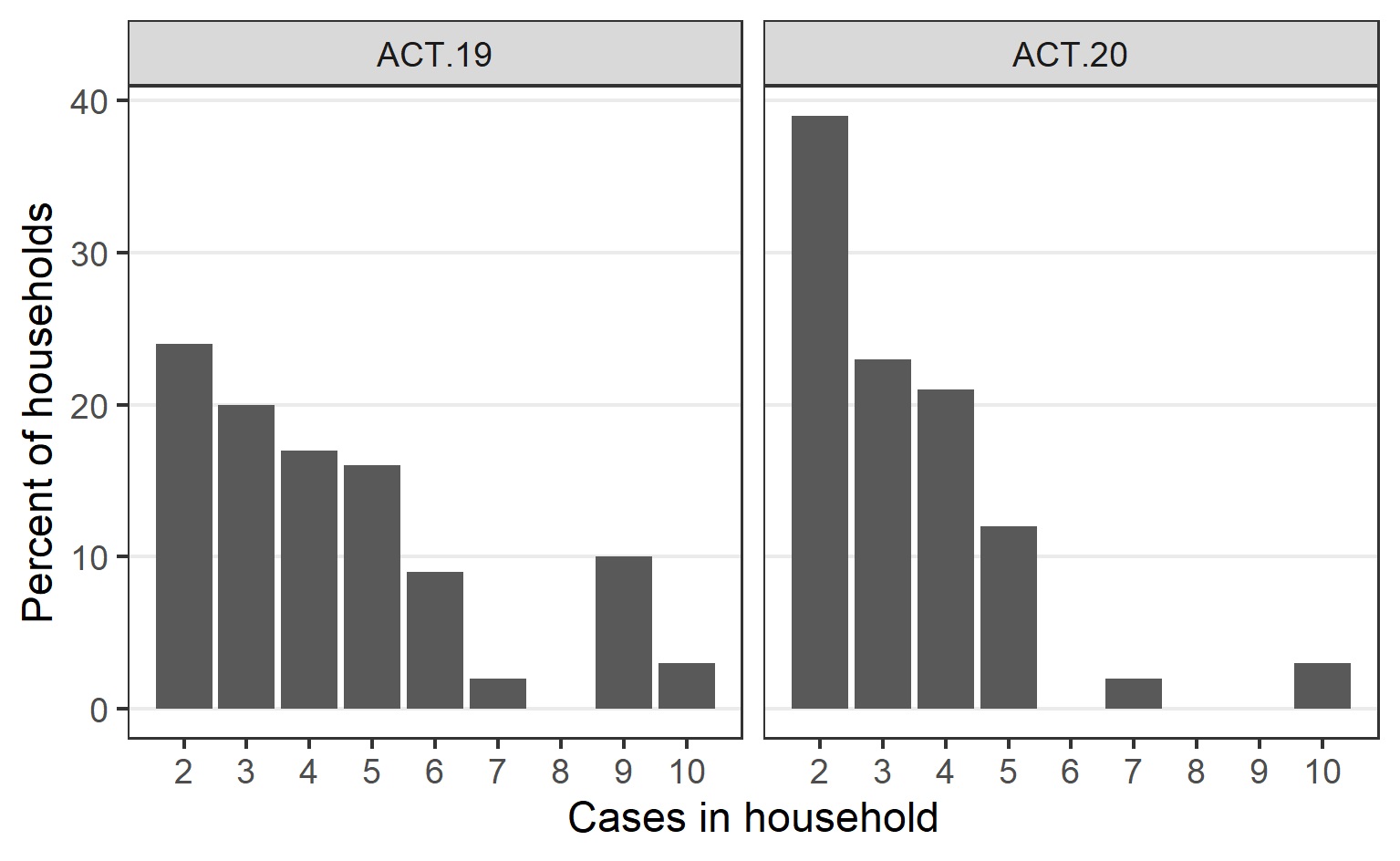
